# The SARS-CoV-2 Integrated Genomic Epidemiology Database (IGED): Linking viral genomes with patient-level metadata to advance statewide genomic surveillance in California

**DOI:** 10.64898/2026.05.14.26353263

**Authors:** Jesse Elder, Rahil Ryder, Mayuri Panditrao, Kaitlin Grosgebauer, Rebecca Katz, Lawrence Tello, Ellaison Carroll, Deva Borthwick, Chaman Kaur, Romario Smith, Victor Shiau, Will Wheeler, Emilia Reilly, Jennifer Myers, Lauren Nelson, Esther Lim, Phacharee Arunleung, Elizabeth Baylis, Sabrina Gilliam, Tamara Hennesy-Burt, Brooke Bregman, Elana Silver, Curtis Kapsak, Sage Wright, Tomas Leon, John Bell, Christina Morales, Debra A. Wadford

**Affiliations:** Viral and Rickettsial Disease Laboratory, Center for Laboratory Sciences, California Department of Public Health, Richmond, CA 94804, USA; Climate Change and Health Equity, California Department of Public Health, Richmond, CA 94804, USA; Informatics Branch, Center for Infectious Disease, California Department of Public Health, Richmond, CA 94804, USA; Public Health Reporting Information Exchange, Division of Communicable Disease Control, California Department of Public Health, Richmond, CA 94804, USA; COVID Control Branch, Center for Infectious Disease, CDPH, Richmond, CA 94804, USA; Theiagen Genomics, Highlands Ranch, CO 80129, USA

**Keywords:** data modernization, genomic data integration, public health data infrastructure, genomic epidemiology, SARS-CoV-2 sequencing

## Abstract

In July 2021, the California Code of Regulations Title 17 required all laboratories performing SARS-CoV-2 whole genome sequencing (WGS) to report their sequencing results to the California Department of Public Health (CDPH). These viral genomic data and patient metadata were compiled into the Integrated Genomic Epidemiology Database (IGED). Linking anonymized viral sequences with patient-level information enabled monitoring of infectiousness, pathogenicity, transmission dynamics, evolution, and vaccine evasion among emerging SARS-CoV-2 lineages. Laboratories performing SARS-CoV-2 WGS transmitted sequencing results to CDPH through Electronic Laboratory Reporting (ELR) and non-ELR pathways. CDPH applied uniform reporting requirements but allowed flexibility in specific data formats to accommodate diverse data systems. To preserve data quality and interoperability across heterogeneous sources, CDPH implemented standardization, validation, and deduplication protocols. Snowflake, a cloud-based data storage and analytics platform, and Posit Connect, a cloud deployment and automation platform, supported the management, processing, and integration of data within the IGED. The IGED established links between SARS-CoV-2 WGS data and epidemiologic metadata for 801,418 sequences, representing 81.7% of all sequences reported in California. Lineages reported to the IGED showed strong concordance with lineage proportions in GISAID. Sequences reported to the IGED had average turnaround times longer than one month, and the majority of sequencing was performed in Southern California and Los Angeles. The IGED enhanced genomic surveillance through predictive modeling and monitoring concerning evolutionary trends such as recombination and saltations in persistent infections. Development of the IGED highlighted the need for standardized data requirements, sustained funding for sequencing, incentives for data submission, and interdisciplinary collaboration to build an effective genomic surveillance system. This framework for linking genomic and epidemiologic data has not only generated critical insights for SARS-CoV-2 but also provided the foundation for CDPH and other public health organizations to develop similar IGED-like systems for other priority pathogens as genomic surveillance expands.

**Author Summary:** In California, the COVID-19 pandemic generated an unprecedented volume of anonymized viral genomic data, creating a critical need to link sequencing results with patient information for genomic epidemiology. To meet this need, we developed the Integrated Genomic Epidemiology Database (IGED), a comprehensive resource that connects SARS-CoV-2 whole-genome sequencing (WGS) data with corresponding patient records. Using cloud-based computational infrastructure, we standardized and integrated submissions from numerous laboratories and jurisdictions, each with distinct technical requirements for providing data to CDPH. Of nearly one million records received, we successfully linked 801,418 WGS records to patient data. The IGED supported public reporting of circulating SARS-CoV-2 lineages, improved understanding of viral evolutionary dynamics, and served as the foundation for a genomic epidemiology tool used in outbreak investigations. By establishing a robust framework for linking WGS and patient-level data, we provide a model that can be adapted by other public health agencies for emerging pathogens of concern.

## Introduction

In March 2020, the Governor of California declared a COVID-19 State of Emergency after which the California Department of Public Health (CDPH) established California COVIDNet, a network of public, private, and academic laboratories to expand SARS-CoV-2 whole genome sequencing (WGS) capacity and output throughout the state (1). Other healthcare systems, academic laboratories, and private laboratories unrelated to California COVIDNet also performed sequencing to meet the emerging need for genomic characterization of the virus. In July 2021, the state of California updated the California Code of Regulations Title 17, Section 2505, subsection (q) to require all laboratories performing SARS-CoV-2 WGS to report their results to CDPH. Title 17 mandated that laboratories report not only the genomic results, but also patient data linked to those sequences.

Although sequence data alone enables broad surveillance of SARS-CoV-2 lineages circulating in California, linking viral genome sequences to patient data is key to gaining deeper insights into transmission dynamics and viral evolution as well as geographic and demographic trends of certain lineages (2, 3, 4, 5, 6). Linking WGS data and patient data is also crucial for monitoring infectiousness, pathogenicity, and vaccine evasion of emerging SARS-CoV-2 lineages (7, 8, 9). Without connections between viral genomic data and patient epidemiologic data, the ability for public health systems to address emerging variants would be curtailed. Moreover, it would become more challenging to assess which emerging lineages pose a greater risk for hospitalization or severe disease.

To ingest and interpret the volume of genomic and epidemiologic data being reported through Title 17, CDPH epidemiologists, data scientists, and genomics experts collaborated to develop a centralized database of Title 17 reportable data. This database linked anonymized WGS data with patient data and was designated the SARS-CoV-2 Integrated Genomic Epidemiology Database (IGED). Here we describe the implementation of the IGED, detail how it has been used for genomic and epidemiologic surveillance of SARS-CoV-2 and provide recommendations for other organizations building similar data systems and expanding them to other pathogens.

## Methods

### 2.1. Implementation of Integrated Genomic Epidemiology Database

Implementing the IGED required establishing a new surveillance framework that did not exist prior to the pandemic that included: 1) creating a new laboratory reporting structure to include SARS-CoV-2 WGS results; 2) onboarding laboratories throughout California to utilize the new reporting structure; 3) developing and automating processes to link sequencing and patient data from multiple sources and integrating them into one dataset; 4) maintaining the centralized dataset by making changes to accommodate data format, lineage, and reporting changes over time; and 5) gleaning public health insights from the dataset to enhance integration of genomic epidemiology into California public health practice. To create, maintain, and utilize the IGED, we collaborated with multiple CDPH teams, California COVIDNet sequencing partner laboratories, and the University of California Santa Cruz (UCSC) Genomics Institute. By September 2024, the IGED was fully automated and actively maintained as a part of routine SARS-CoV-2 genomic surveillance. This database continues to be used for CDPH analysis, tools, dashboards, and to inform public health action and decisions pertaining to SARS-CoV-2.

### 2.2. Laboratory Reporting Structure for SARS-CoV-2 Whole Genome Sequences

Title 17 established data reporting requirements for laboratories performing SARS-CoV-2 molecular tests and WGS in California. Laboratories were required to submit electronic laboratory reports (ELR) for each COVID-19 test and whole genome sequence to CDPH via Health Level Seven (HL7) message or comma separated values (CSV) file. These reports were then ingested into the State of California’s electronic disease reporting and surveillance system, the California Reportable Disease Information Exchange (CalREDIE) (https://www.cdph.ca.gov/Programs/CID/DCDC/Pages/CalREDIE.aspx).

Laboratories reported ELRs using several Logical Observation Identifiers Names and Codes (LOINC) containing WGS metadata, namely SARS-CoV-2 subspecies classifications and unique identifiers. In particular, SARS-CoV-2 Pango Lineages were most commonly reported with the 96741-4 LOINC, but the codes 96895-8 and 100157-7 were also allowable (Table 1). The Pango lineage system is widely recognized as the standard for determining lineages of SARS-CoV-2 (10). CDPH also collected reports for NextClade Clades (11) using the codes 96896-6 and 94764-8. Additionally, laboratories that performed WGS were required to send a follow-up ELR containing unique sequence identifiers using the GISAID accession (96766-1) or Sequence Study ID, also called a sample ID, (98062-3) LOINC codes. All messages originating from a single test contained the same core data fields including but not limited to the accession number and collection date of the original diagnostic test, the name of the testing and sequencing facilities, and patient Personally Identifiable Information (PII) data fields such as patient name, gender identity, race and ethnicity, county of residence, and date of birth.

**Table 1.**
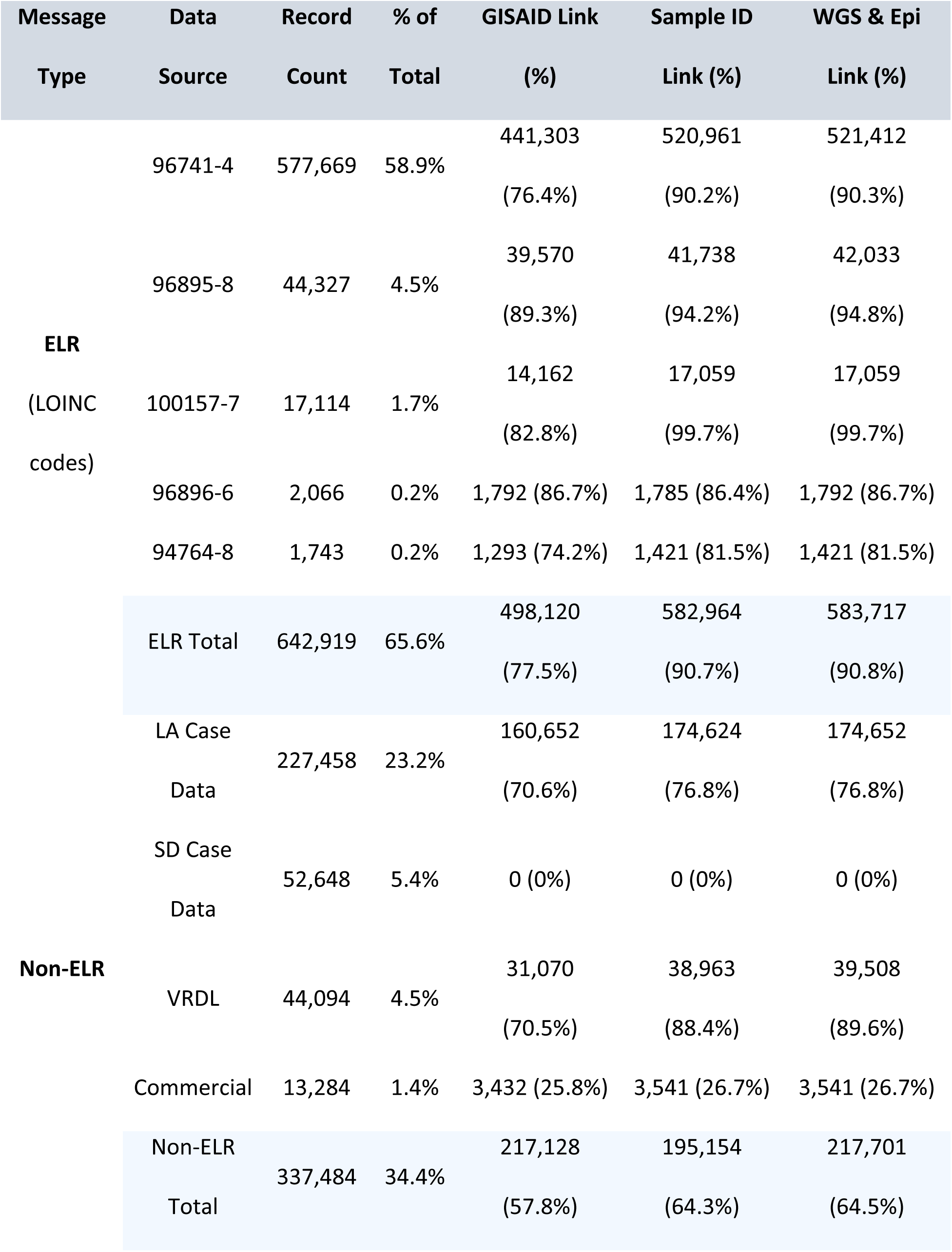
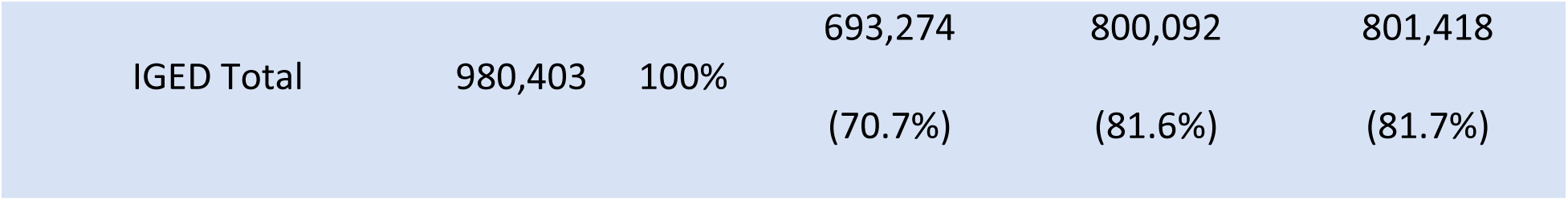
Integrated Genomic Epidemiology Database Sources and Linkages.

### 2.3. Establishing WGS Reporting Across California

CDPH leveraged existing relationships with laboratory partners to implement WGS data reporting in response to the evolving COVID-19 pandemic. At the outset of the pandemic, WGS reporting was relatively new to public health practice, therefore laboratory personnel across the state were trained how to transmit these reports with the correct data fields and in the proper format. Many laboratories also needed to develop systems to join their generally separate WGS and patient data systems to generate and transmit WGS reports. To provide flexibility in methods of reporting while also maintaining compatibility with CalREDIE structure, CDPH adapted existing workflows to allow for secure reporting of highly structured CSV files in addition to standard HL7 messages to CalREDIE. Extensive testing was performed on CSV and HL7 messages to ensure that messages were formatted properly for their LOINC codes and Systematized Nomenclature of Medicine – Clinical Terminology (SNOMED) codes prior to importing into CalREDIE. This included checking the quality of the reported Pango Lineages, Sequence IDs, and collection dates as well as patient demographic data used to match to CalREDIE records. Due to the rapid initial response needed to address the pandemic, we provided laboratories with other non-ELR routes to securely transmit WGS reports which did not meet the strict structural requirements of ELRs and the CalREDIE database. These non-ELR transmission methods included secure emails of loosely structured data from commercial laboratories, better defined datasets internal to the CDPH Viral & Rickettsial Disease Laboratory (VRDL), and richer case reporting datasets from Los Angeles and San Diego counties, both of which maintain separate disease reporting systems with similar structure to CalREDIE. Non-ELR methods were intended as a stopgap measure to eventually be discontinued once laboratories onboarded to CalREDIE WGS reporting.

### 2.4. Expanding Network of Laboratories Reporting WGS Results

We dedicated significant resources to onboarding partner laboratories for SARS-CoV-2 WGS reporting. Because each facility managed data through different LIMS vendors, we provided tailored support to navigate varying technical capabilities. While some laboratories were more prepared to report WGS results, others required direct CDPH intervention and guidance to meet reporting standards. Certain public health laboratories (PHLs) and hospitals already used HL7 messaging standards which expedited the transition to WGS reporting to CalREDIE. Conversely, some academic laboratories lacked reporting infrastructure and required extensive assistance to ensure compliance with reporting guidelines.

To enhance data quality and interoperability, we expanded the WGS metadata requirements beyond Pango lineage information. It was essential to report sequence identifiers, such as GISAID accession numbers and sample IDs, to link patient data directly with sequences in public repositories. Some commercial laboratories, such as those contracted by CDC, already provided these identifiers to CDPH via HL7 messages or an existing Secure File Transfer Protocol (SFTP). In many cases, reporting these additional fields was a straightforward change for laboratories as it only required replacing the Pango Lineage information with sequencing identifiers in otherwise largely identical ELRs. However, certain resource-limited laboratories, namely local PHLs, required more flexibility. For these partners, we leveraged a partnership with Theiagen Genomics (Highlands Ranch, CO United States) to facilitate the sharing of deidentified unique accession numbers through California COVIDNet Terra using the Terra.bio platform (https://terra.bio). This allowed CDPH to securely link the genomic data to internal patient records while minimizing the reporting burden on PHLs.

### 2.5. Centralized Repository for Genomic and Epidemiological Data

A key innovation of the IGED was linking patient data in the CDPH data ecosystem with SARS-CoV-2 WGS data for the purpose of genomic epidemiology. The IGED serves as a centralized repository consolidating viral WGS results from both ELR and non-ELR sources. Each record links WGS metadata such as Pango lineages, NextClade Clades, and unique sequence identifiers to patient data such as demographics, addresses, and unique disease incident identifiers. These direct linkages between patient and WGS data have enabled outbreak investigations (https://pathogengenomics.ucsc.edu/tools), surveillance of concerning patterns of viral evolution (12, 13), and comparisons of clinical outcomes for patients infected by viruses with different lineages and mutations (14, 15). A summary of the data flow contributing to the IGED and to overall genomic surveillance at CDPH is provided in Figure 1.

**Fig 1.**
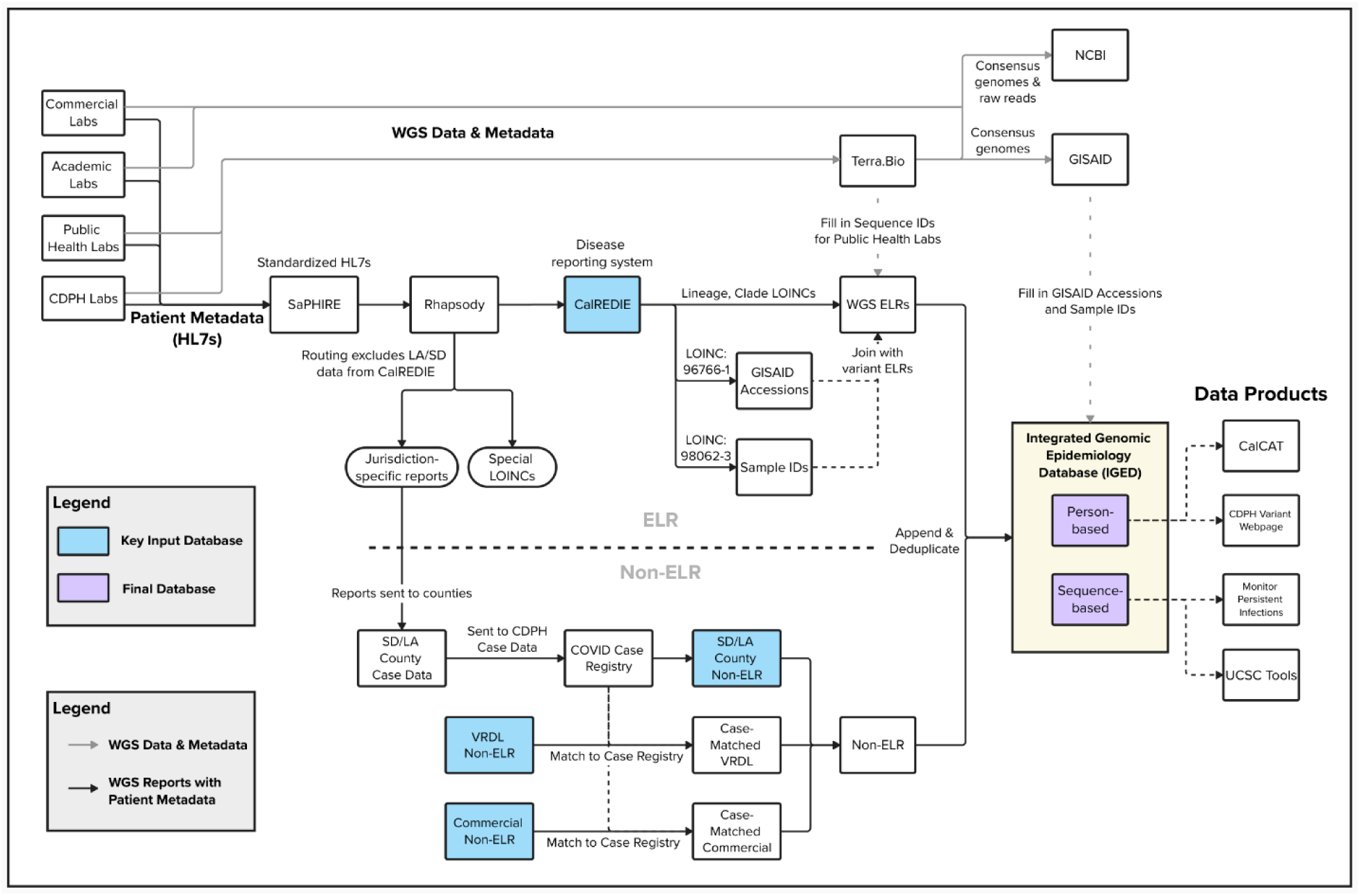
Integrated Genomic Epidemiology Database (IGED) Dataflow. Blue boxes represent the ELR input data source, CalREDIE, and the four non-ELR input data sources: Los Angeles, San Diego, VRDL, and Commercial laboratories. Purple boxes indicate the two versions of the IGED with per person (“person-based”) and per sequence (“sequence-based”) deduplication.

### 2.6. Developing & Maintaining the IGED

#### 2.6.1. Data Management Across Laboratories and Jurisdictions

The expansive network of laboratories and their disparate WGS reporting methods posed several challenges to creating a homogeneous dataset suitable for integrating reports across all local health jurisdictions and laboratories. While most laboratories submitted ELRs to CDPH, not all WGS reports could be incorporated into CalREDIE. Laboratories performing sequencing in Los Angeles and San Diego counties would report WGS ELRs to CDPH, but those reports were diverted away from CalREDIE using the Rhapsody (https://rhapsody.health/) system due to separate disease reporting systems in those counties (Fig 1). WGS reports from Los Angeles and San Diego counties were instead reported through separate non-ELR mechanisms which led to structural inconsistencies between data sources. We conducted extensive data mapping to find matching data fields between ELR and non-ELR data sources, but several CalREDIE-specific fields remained unavailable in non-ELR. To ensure the interoperability of the IGED, we prioritized the standardization of core fields outlined in CDPH guidelines for Title 17.

#### 2.6.2. Data Standardization

We employed standardized data-processing pipelines to harmonize SARS-CoV-2 lineages and metadata across the IGED. Although laboratories were required to report Pango Lineages following the strictly formatted Pango nomenclature, many reports contained data artefacts such as special characters, usually due to technical differences in the Laboratory Information Management System (LIMS) of each laboratory, or due to formatting inconsistencies and data entry errors that were more common in non-ELR data sources. To ensure consistency, we conducted extensive data cleaning to remove these artefacts and correct formatting issues. We then found exact matches between the standardized reported lineages and an updated record of the Pango nomenclature. For reports without an exact match, the IGED instead used *fuzzy regex* joins with the R package fuzzyjoin v0.1.6 (16) to find the closest match to the Pango nomenclature.

To standardize PII, we removed non-word characters, white spaces, middle names, and prefixes from patient names. All date fields, including date of birth and specimen collection date were set to the international standard YYYY-MM-DD date format. For some non-ELR reports, additional steps needed to be taken to ensure completeness and accuracy of the patient epidemiologic data. Although some non-ELR sequences contained limited patient data, comprehensive patient data were available in the COVID-19 Case Registry (Fig 1). To find matching records in the Case Registry, we used the RecordLinkage v0.4-12.1 R package (17). First, records were matched on standardized date of birth, then the similarity of first and last names were calculated using the Jarowinkler method. Records with an EpiLink match score >94% were kept as likely matches (17). This process also standardized patient names to match CDPH records to facilitate downstream deduplication.

#### 2.6.3. Deduplication of WGS Reports: Person vs Sequence Based Databases

While record duplication was rare, laboratories occasionally reported the same sequence multiple times, and these duplicates had to be filtered out. In some duplication cases, laboratories reported corrections to their original WGS ELRs, while in other cases, laboratories reported both Pango Lineages and Clades for the same sequence. Duplication between data sources also occurred. Some sequences initially reported via non-ELR were reported again via ELR to CalREDIE. ELR records were standardized and had more complete data fields, so we prioritized keeping these records during deduplication.

To address epidemiological analysis needs, two versions of the IGED database were created with different deduplication schemes: a person-based database referred to as the IGED, and a sequence-based database designated as IGED_SEQ (Fig 1). The person-based scheme removed duplicate records based on first name, last name, date of birth, and viral lineage in order to extract one sequence per person per infection. All results in this article are based on the person-based IGED. The sequence-based scheme additionally deduplicated on collection date and aimed to keep one record per sequence regardless of the number of sequences per person. This deduplication scheme was used to monitor all sequences reported to CDPH while still minimizing record duplication. Having a comprehensive, sequence-based database of WGS in California enabled the characterization of within-host viral dynamics (13) and outbreak investigations (https://pathogengenomics.ucsc.edu/tools/cluster-tracker).

### 2.7. Maintaining Connections Between Databases

#### 2.7.1. Integrating Case Data Systems

The IGED incorporated data from three separate case reporting systems: CalREDIE, Los Angeles (LA) Case Data, and San Diego (SD) Case Data. These databases used unique incident identifiers to correspond to one period of illness for each patient, but the identifier formats were different between the three systems. To improve interoperability of these systems, we maintained the California COVID-19 Case Registry that consolidated unique incident identifiers from all three systems into one database. Since CalREDIE records were accessible to CDPH, we specifically extracted WGS-related records for LA and SD from the Case Registry to populate the IGED (Fig 1). Consolidating the three systems into the IGED ultimately allowed us to establish a statewide system linking the anonymized SARS-CoV-2 WGS data and patient epidemiologic data.

#### 2.7.2. Connecting Genomic and Epidemiological Data

The IGED linked extensive patient data in the CDPH data ecosystem with WGS data stored in separate sequencing databases. This linkage relied on two primary unique identifiers: sample IDs and GISAID accession numbers. Accurate data linkage required sequence identifiers to be unique, unambiguous, and consistent with identifiers in GISAID, GenBank, and California COVIDNet Terra. However, sample IDs in the IGED often contained additional prefixes and suffixes inconsistent with other databases. In order to standardize sample IDs to match WGS databases, we established definitive formatting rules that prioritized brevity while including the identity of the sequencing laboratory or specific surveillance effort showcased with the Patient Anonymized Unique Identifier (PAUI) system implemented by California COVIDNet (1). Our formatting rules aimed to remove extraneous data and get to the “root” of sample identifiers such that they would be easily recognizable to the sequencing laboratories.

Maintaining consistent formatting between databases enabled the IGED to routinely join to and pull from the GISAID database (Fig 1). The GISAID database routinely updates Pangolin Lineage calls. GISAID also contains both sample IDs and GISAID accession numbers for every sequence, whereas ELRs from CalREDIE generally had either one or the other. Therefore, by leveraging the connection to GISAID, the IGED was able to maintain up-to-date lineage calls and fill in both sample IDs and GISAID accession numbers for all matching records. Additionally, IGED records with a sample ID but no GISAID accession enabled connections to other WGS databases like GenBank and California COVIDNet Terra.

### 2.8. Data Management: Processing, Analysis, and Storage

In response to the increasing storage and computational demands posed by the unprecedented volume of COVID-19 data, CDPH migrated all COVID-19 surveillance data, including WGS reporting data, to the Integrated Infectious Disease Data Warehouse (I2D2), a cloud-based data warehouse hosted by Snowflake (Bozeman, MT, United States) for the storage, processing, and rights-based access control to these sensitive public health data. SARS-CoV-2 WGS ELRs were imported from CalREDIE into I2D2, whereas non-ELR data were sent through SFTP, stored on CDPH network drives, and then loaded into I2D2. IGED data processing scripts were built using R code in Posit Workbench and SQL code in Snowflake Views and stored procedures. These scripts were then automated using Posit Connect servers (Boston, MA, United States) and scheduled tasks in Snowflake. Larger queries were run using Snowflake’s cloud compute resources while smaller intermediary datasets were pulled into Posit Workbench for computing on POSIT servers. Code run from RStudio and Posit Workbench (18) used odbc v1.3.2 (19) to connect to and pull data from Snowflake. Both intermediary and final output tables were stored in I2D2. We identified new and changed records in each IGED data source every week which allowed for incremental processing, reducing the need to reprocess observations wherever possible and minimizing compute costs and processing time. The IGED data pipeline incorporated logging and error handling to ensure processing of each data source was successful prior to combining them into the IGED.

### 2.9. IGED Data Quality Monitoring

To ensure the accuracy of ELR and non-ELR records in the IGED, CDPH used data quality metrics and dashboards to monitor for data discrepancies. After completion of the weekly IGED data pipeline update, automated emails were sent to data managers summarizing total IGED sequence counts by month and by reporting method (ELR, LA Case Data, SD Case Data, Commercial non-ELR, and CDPH non-ELR). These summaries were provided for the IGED in the current week as well as for previous versions of the IGED in past weeks. This allowed for rapid detection of processing errors related to integrating the various input databases.

Additionally, we created a data quality dashboard in Snowflake to automatically generate graphs and metrics for detection of reporting inconsistencies. For example, comparing total sequence counts with counts of sequences that had a GISAID ID and a Sample ID (% with WGS & Epidemiology Link, Table 1) gave an overview of how many patient records were connected with WGS data in public repositories. This dashboard also tracked the number of reports with valid, known Pango lineages over time. This allowed us to identify laboratories submitting improperly formatted WGS reports as well as laboratories with low sequencing success rates. Other uses for this dashboard included tracking where sequences originated to identify areas that were not proportionately represented by WGS data, highlighting inequities in sequencing efforts.

### 2.10. Data Security

The IGED contained both PII and PHI (Protected Health Information) and therefore data security measures were necessary and required. Lineage data, PII, and PHI were stored securely in I2D2 using role-based access controls (RBAC), which is a practice that assigns groupings of permissions to roles and then assigns those roles to users. This method of grouping access makes it easier to monitor, control, and maintain data governance measures at scale. CDPH data managers created Snowflake roles specific to COVID-19 WGS data and permitted an authorized list of users to access these data.

## Results

### 3.1 Overview of the IGED

The primary objective of the IGED was to link genomic data with patient data to inform targeted and data-driven public health policies and decisions. Through collaboration with many CDPH and local partners, we successfully integrated genomic and epidemiological data for SARS-CoV-2 with the IGED. Laboratories reported WGS results such as Pango Lineages, NextClade Clades, and sequence identifiers through CalREDIE ELR messages and other non-ELR sources. The IGED ingested data from seven different LOINC codes, five of which included viral classifications, as well as four different non-ELR data sources. Over 50 laboratories in California including commercial, academic, state, and local PHLs contributed to data in the IGED. The IGED includes sequences from California COVIDNet laboratories (1) including 14 of the 29 PHLs in California, and laboratories outside of California COVIDNet such as Kaiser Permanente SCPMG Regional Reference Laboratory and Aegis Sciences sequencing for Walgreens, among others. As of February 2026, the IGED was being actively maintained as part of routine SARS-CoV-2 genomic surveillance.

### 3.2 Data Integration and Connectivity

As of April 2025, 980,403 sequences had been reported to the IGED; 877,583 (89.5%) included a successful lineage call and 801,418 (81.7%) had a direct link between genomic and epidemiological data via a sequence identifier (Table 1). The majority of sequences were reported via ELR (65.6%) while 34.4% of sequences were reported via non-ELR, most of which came from Los Angeles County (23.2%). As shown in Table 1, most but not all records with a WGS & Epi Link (81.7%), either a GISAID accession or a sample ID, contained a sample ID (81.6%). Fewer records overall linked directly to GISAID (70.7%) due to sequences being rejected by or not submitted to GISAID. ELR data were much more likely to contain GISAID accessions (77.5%) and sample IDs (90.7%) than non-ELR data (57.8%, 64.3%). WGS & Epi linkage rates from commercial laboratories (26.7%) and San Diego (0%) non-ELR data were particularly low. Commercial laboratory non-ELR was used early in the pandemic prior to most WGS data reporting standards, so sequence identifiers were reported infrequently, whereas San Diego did not report any sequence identifiers throughout the pandemic.

The number of records and the percent of the total records in the IGED per ELR data source (LOINC codes) and per non-ELR data source as of April 2025. Overall number and percentage of records in each Message Type (ELR or Non-ELR) and the entire IGED are highlighted in blue. GISAID Link, Sample ID Link, and WGS & Epidemiology Link columns show the number of records that contain a GISAID accession, a Sample ID, and either a GISAID accession or a Sample ID, respectively. The percentages for GISAID Link, Sample ID Link, and WGS & Epi Link are calculated as the number of records per Data Source in these columns divided by the total Record Count per Data Source.

### 3.3 Sequencing Results and Timeliness

In addition to directly linking 693,274 sequences in GISAID to patient data in the IGED (Table 1), the lineages reported to the IGED, including those without a GISAID link, were largely similar to the GISAID database. Figure 2A shows the proportion of sequences assigned each major lineage for all sequences in the IGED and GISAID with collection dates between March 2020 and December 2024 aggregated quarterly. In order to compare major lineage proportions between the two databases, we used a two-tailed Spearman correlation test implemented in *cor.test* of the R stats package (20). Across the 21 quarters analyzed, 19 demonstrated a strong and statistically significant correlation between the ranking of major lineages in the IGED and in GISAID (Spearman ρ = 0.643–0.986; S1 Table), indicating that lineage proportions were largely consistent between the two databases over time. Lower correlations occurred during periods when closely related Omicron lineages, such as B.1.1.529, BA.1, and BA.1.1, were predominant.

**Fig 2.**
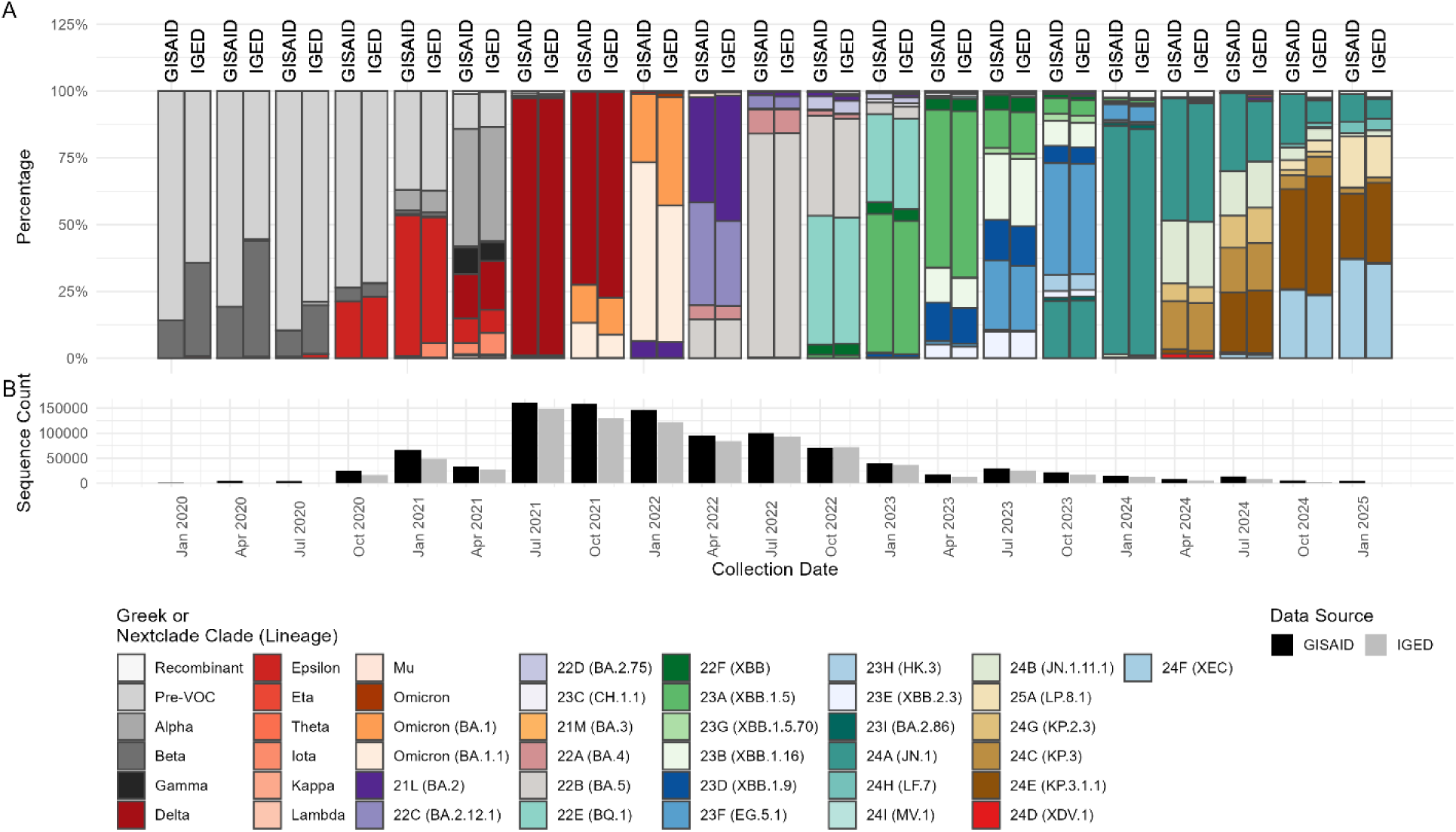
IGED and GISAID SARS-CoV-2 Sequence and Lineage Comparison Over Time. **A.** Comparison of California SARS-CoV-2 variant World Health Organization (WHO) Greek designations and Nextclade Clade proportions in California between the GISAID repository and the person-based IGED graphed per quarter including data from March 2020 through the end of February 2025. SARS-CoV-2 variant WHO Greek designations and Nextclade Clades were assigned to each reported Pango Lineage. **B.** Number of California SARS-CoV-2 sequences reported to each database over time. The X-axis is the collection date shown in month and year and grouped into three-month time periods. Figure was created with GISAID and IGED data using the ggplot2 v3.5.2, RColorBrewer v1.1.3 and patchwork v1.3.2 packages in RStudio v4.4.1 (20, 21, 22, 23).

This may be because many sequences reported to the IGED did not link to GISAID and therefore had static lineages that did not update with the lineages in GISAID. Additionally, the first two quarters of 2020 did not show significant correlations – likely due to the low numbers of sequences reported to the IGED during these periods (Fig 2B, S1 Table). Overall, IGED sequence volumes tended to be lower than the number of sequences in GISAID and particularly prior to the changes to Title 17 implemented in July 2021 (Fig 2B). Moreover, certain sequencing laboratories either did not have resources to send WGS reports to CDPH or did not collect patient data for their sequences and were therefore unable to report, which may explain the differences in overall sequencing volumes.

Additionally, we monitored sequencing turnaround time for each sample. This was calculated as the number of days between the earliest date associated with a sample (i.e. positive PCR test) and the date the sequence was reported to CDPH (Fig 3). A majority of the sequencing facilities were part of California COVIDNet (1) and many, but not all, of the commercial laboratories were contracted by the US Centers for Disease Control and Prevention (CDC) (24, 25, 26). Commercial sequencing facilities averaged the shortest turnaround time of 21.7 days. Academic, local PHLs, and CDPH sequencing facilities averaged 65.1, 73.9, and 63.2 days, respectively. However, higher averages and peaks in Figure 3 may be skewed by sequencing historical samples and reporting backlogs due to the onboarding process for new sequencing laboratories.

**Fig 3.**
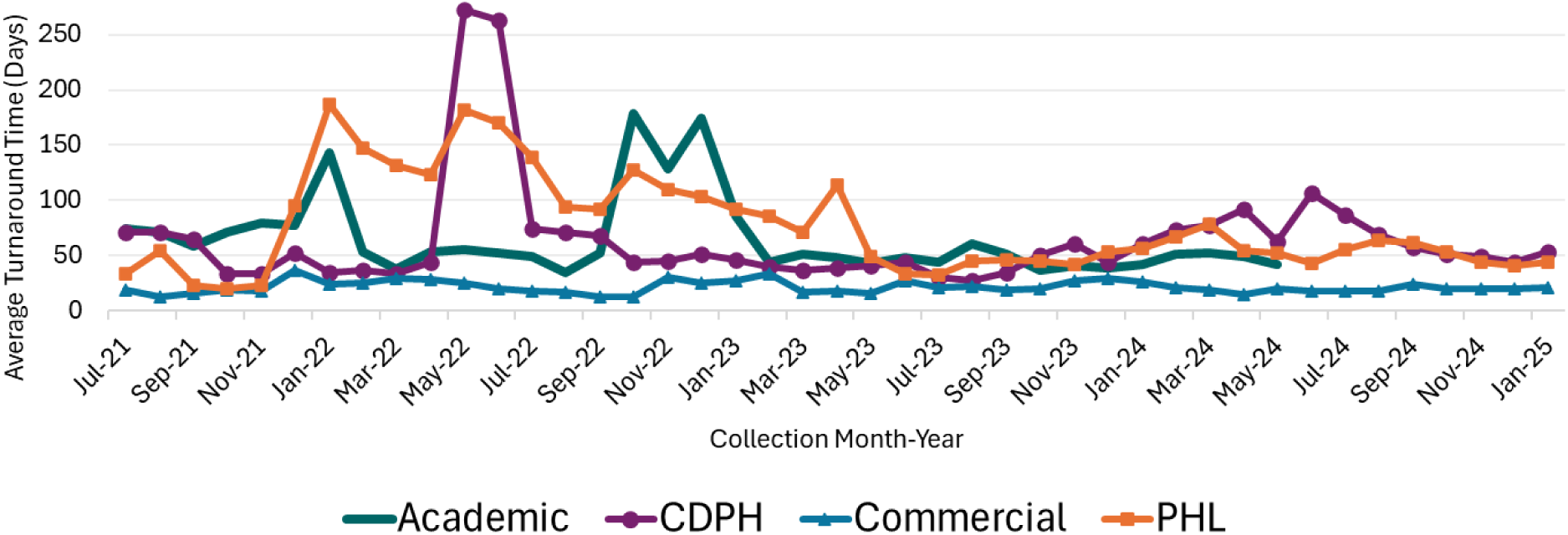
Average Turnaround Time of WGS Reporting Per Sequencing Facility Type. Average turnaround time (days) from the earliest date associated with a sample to when the sequence was reported to CDPH over time per sequencing facility type. Sequencing facilities include academic laboratories, California Department of Public Health (CDPH), local public health laboratories (PHL), and commercial laboratories. The majority of commercial laboratories were contracted by the CDC. The vast majority of non-ELR records have an unknown turnaround time and therefore only ELR-reported sequences are shown.

### 3.4 Sequencing Representativeness

The types of laboratories that performed sequencing and reported results to CDPH varied substantially by geographic region (Fig 4). The Southern California and Los Angeles Regional Public Health Officer (RPHO) Regions contributed 32.7% and 26.7% of California SARS-CoV-2 sequencing, respectively, primarily performed by commercial laboratories. The Bay Area RPHO region contributed 21.0% of California SARS-CoV-2 sequences with the largest proportion contributed by local PHLs compared to the rest of the state. Central California, Greater Sierra Sacramento, and Rural North RPHO regions contributed 10.7%, 7.0%, and 1.6%, respectively by a variety of sequencing facilities. The Rural North RPHO region relied heavily on California COVIDNet and academic laboratories, many of which were also contracted by California COVIDNet.

**Fig 4.**
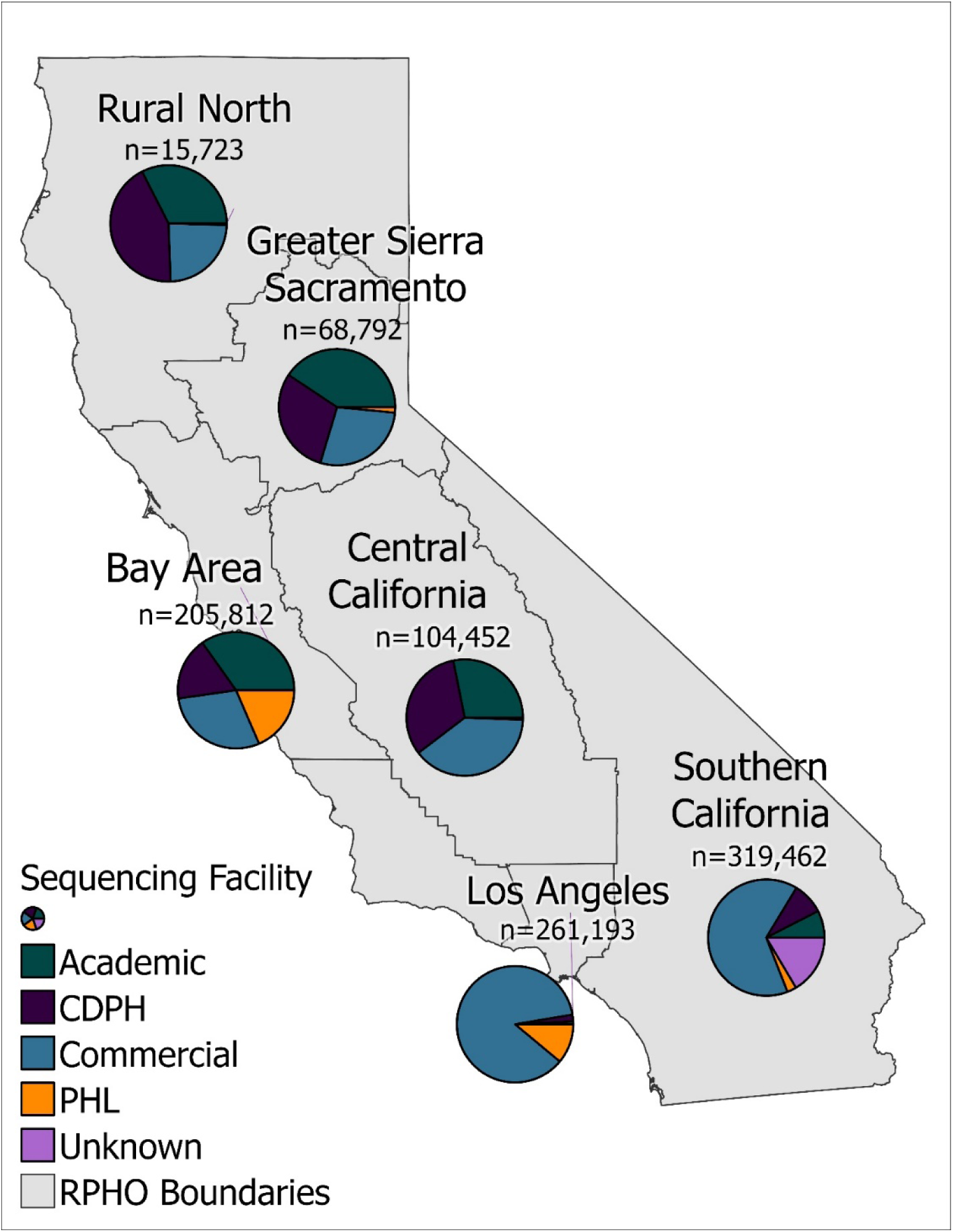
SARS-CoV-2 Sequence Counts Per California Public Health Officer Region by Sequencing Facility. Regional Public Health Officer (RPHO) boundaries with sequencing facility type proportions displayed via pie chart. Sequencing facilities include academic laboratories, California Department of Public Health (CDPH), local public health laboratories (PHLs), and commercial laboratories, some of which were contracted by the CDC. The n value is the total sequencing counts from the Integrated Genomic Epidemiology Database (IGED) per RPHO region as of April 2025. Map was created using ArcGIS Pro (version 3.0.0). The original county lines were downloaded from https://catalog.data.gov/dataset/ca-geographic-boundaries. This is an open access dataset in the public domain and no license information is provided.

### 3.5 Enhancing Genomic Surveillance with the IGED

CDPH used the IGED to monitor SARS-CoV-2 lineages over time, inform public health policies and decisions, and take relevant public health actions. The IGED’s SARS-CoV-2 lineage proportions were regularly updated on the CDPH Variant Webpage (27) and were used to forecast lineage proportions using the California Communicable diseases Assessment Tool (CalCAT) (28, 29).

Although the IGED generally had fewer sequences than other sequencing repositories, the patient data in the IGED facilitated genomic epidemiology investigations. The IGED was the primary source of patient data for the Big Tree Investigator tool (https://pathogengenomics.ucsc.edu/tools) which automatically overlayed patient data onto phylogenetic trees generated by UShER (30), streamlining the detection and investigation of outbreaks. Additionally, the IGED contained unique identifiers (Incident ID) and PII (First Name, Last Name, Date of Birth) that allowed for linkage to other CDPH databases such as the COVID-19 Case Registry, COVID-19 Hospitalization Registry, and the Vaccine Registry further enhancing data interconnectivity and improving public health decision making.

## 4. Discussion

### 4.1 Public Health Impact and Accomplishments

From nearly one million reported California SARS-CoV-2 whole genome sequences (WGS), we successfully linked 801,418 (81.7%) anonymized SARS-CoV-2 sequence data with patient epidemiologic data (Table 1) and catalogued them in an integrated database, the Integrated Genomic Epidemiology Database (IGED). The IGED contains standardized SARS-CoV-2 WGS results from numerous jurisdictions, laboratories, and databases throughout California. Here, we have established a framework for reporting SARS-CoV-2 WGS data linked to patient information. As genomic surveillance becomes more routinely utilized, this framework can be adapted for other pathogens of public health significance.

The IGED streamlined access to patient data otherwise unavailable or resource-intensive to acquire with only anonymized WGS data. The IGED’s connection to the UCSC Big Tree Investigator tool allowed for overlaying epidemiologic data directly onto a phylogenetic tree, quickly verifying relatedness between samples. This tool effectively identifies genomically related sequences, which can be indicative of outbreaks that were not initially captured by targeted sequencing efforts. Furthermore, using the IGED, CDPH was able to proactively inform local health jurisdictions about circulating lineages of concern including those associated with increased disease severity or transmissibility (31), those arising from ongoing persistent infections (13), or those possessing mutations that conferred resistance to therapeutic interventions (32).

Linking SARS-CoV-2 sequence data with patient epidemiologic data allowed for meaningful and actionable public health surveillance as the pandemic proceeded. Using the IGED, we successfully identified patients with persistent SARS-CoV-2 infections which were disproportionally male and observed selective pressures and convergent evolution of mutations acquired within host (13). Epidemiologic characteristics of recombinant SARS-CoV-2 lineages within California were published where an increase in viral recombination of SARS-CoV-2 over time was identified and is expected to increase as the virus diversifies (12). The IGED could also be used to gain similar insights into the genomic and epidemiologic factors contributing to reinfections. The IGED also allows for higher data resolution, when needed, for outbreak investigations and/or insight into the viral genomics underpinning severe cases of COVID-19.

Although GISAID had a higher number of sequences than the IGED (Fig 2B), the IGED had more comprehensive patient-related epidemiologic data which allowed for downstream public health applications as discussed above. Moreover, the lineages reported to the IGED strongly correlated with those in GISAID over time. As a result, we relied on the IGED for updating SARS-CoV-2 lineage proportions and SARS-CoV-2 lineage predictions for public consumption via the CDPH Variant Webpage (27) and the California COVID Assessment Tool (CalCAT) (28, 29), respectively. Understanding the lineage proportions specific to California informed public health actions. For example, CDPH informed healthcare providers to prioritize the use of antivirals, Paxlovid and remdesivir, for treatment of mild to moderate COVID-19 when Omicron became the dominant lineage in California, as certain monoclonal antibodies, REGEN-COV™ and bamlanivimab with etesevimab, were found to be ineffective against Omicron sublineages (32). Continued genomic surveillance is essential to assess effectiveness of vaccines and therapeutics.

### 4.2 Challenges

The primary challenge in development of the IGED was consolidating WGS reports from numerous jurisdictions and laboratories into a centralized, standardized database. Although Title 17 provided clear guidelines and mandates for reporting WGS data, differences in laboratory resources, case reporting systems, data formatting, and data availability hindered IGED development.

In order to conduct WGS, laboratories needed trained personnel capable of performing data entry, sample extractions and sequencing, bioinformatic analysis, and uploading sequences to public repositories. Laboratories also needed significant LIMS expertise to link WGS results with patient data and then securely transmit the data to CDPH via HL7 message. In many cases CDPH provided extra guidance to laboratories about the content and format of the HL7 messages, but some lesser-resourced laboratories lacked the necessary staff to report their data. Even in laboratories with reporting capabilities, CDPH had few mechanisms to enforce Title 17 WGS reporting mandates which led to slower uptake overall.

Combining WGS reports from Los Angeles and San Diego case reporting systems with CalREDIE WGS ELRs required extensive data standardization efforts. Many data fields present in CalREDIE were formatted very differently in these counties’ databases. For example, lineages in the county databases often lacked period delimiters usually present in the highly standardized Pango Lineage nomenclature, which required us to perform fuzzy matching to find the closest matching lineage. Initially, sequencing identifiers and other key fields needed to link WGS and patient data were not available at all in the county databases. Los Angeles eventually reported these fields after CDPH provided guidance on improving their WGS reporting and integration systems. However, San Diego was unable to report these fields primarily due to the San Diego Epidemiology and Research for COVID Health (SEARCH) Alliance sequencing laboratories (https://searchcovid.info/) providing the county with only anonymized WGS results and therefore had zero linkage in the IGED.

Ensuring that sequence identifiers were reported, in the proper format, and could accurately link to sequence data in WGS repositories was a consistent challenge. Some laboratories that sent initial lineage reports were unable to send additional sequence ID reports again due to staffing and resource constraints. To accommodate the needs of different laboratories, the IGED incorporated sequence identifiers from a variety of sources including non-standard HL7 fields of lineage reports, lab-specific SFTPs, and the California COVIDNet Terra database. Sometimes reported sample IDs did not match to a sequence in GISAID due to GISAID altering the IDs after submission. In other cases, separate sequence naming schemes were used for California COVIDNet Terra, GISAID, and GenBank, and only the IDs from one of those databases were reported to the IGED.

### 4.3 Limitations

The IGED faced several limitations in terms of data quality, representativeness, and timeliness. Some of these limitations were specific to the COVID-19 emergency response, but others may also be applicable to future genomic surveillance systems. At the outset of the pandemic, it was a challenge to onboard laboratories to report WGS results because they lacked modern data systems and trained personnel to submit such reports. While CDPH invested in the development of ELR and disease reporting infrastructure, non-ELR reporting methods were also accepted which resulted in more missing data, nonstandard formatting, and data entry errors. These data quality issues sometimes required the use of fuzzy matching of lineages and led to incorrect lineage assignments. Although numerous laboratories now have the capacity to report WGS ELRs to CDPH, during the development and implementation of the IGED the lack of such infrastructure hindered accurate and timely reporting of SARS-CoV-2 WGS results.

Other limitations of the IGED are surveillance bias and geographical representativeness. The IGED was unable to delineate between baseline surveillance samples and targeted investigations in high-risk settings or outbreaks. These targeted samples are likely overrepresented in the IGED. Certain laboratories and time periods may also be overrepresented as a result of duplication between ELR and non-ELR data sources. Future surveillance systems should endeavor to include data tags indicating baseline surveillance and to limit the inclusion of semi-structured non-ELR data in order to minimize surveillance bias. Due to the resources needed to perform sequencing and reporting, Rural North and Greater Sierra Sacramento Regional Public Health Officer regions were underrepresented while Los Angeles was overrepresented (Fig 4).

Turnaround times for reported WGS data remained a consistent limitation throughout the pandemic (Fig 3). CDPH, local PHLs, and academic laboratories generally exhibited longer turnaround times than commercial laboratories, largely due to logistical factors such as specimen shipping, processing, and distribution across the California COVIDNet laboratory network. CDPH also faced delays associated with reporting backlogs, particularly during periods when new sequencing laboratories were being onboarded. In contrast, commercial laboratories achieved the shortest turnaround times, likely because they had onsite access to in-house SARS-CoV-2–positive specimens and contractual obligations to CDC.

To help bridge this timing gap in surveillance, the CalCAT nowcasting model (28, 29) was developed; however, delays continued to constrain the timeliness of genomic epidemiology investigations, including assessments of concerning lineages and clusters identified through tools such as the Big Tree Investigator. While historical surveillance data remain essential for understanding and preventing future outbreaks, reducing turnaround times is critical for enabling real-time genomic investigations outside of known outbreak settings. For active outbreak response, close coordination between genomic epidemiologists and laboratory partners is particularly important to accelerate sequencing workflows.

Sequence repositories such as GISAID and California COVIDNet Terra often detected emerging lineages earlier than the IGED because of lags associated with ELR reporting. To address this discrepancy, CDPH implemented automated alerts in Looker (https://cloud.google.com/looker) based on California COVIDNet Terra data, enabling earlier detection of concerning lineages and mutations and mitigating delays caused by IGED data-matching processes.

### 4.4 Recommendations and Conclusions

The COVID-19 pandemic revolutionized genomic surveillance making it a worldwide endeavor to advance knowledge about the virus and its evolution, to inform public health action and decisions, and assess effective therapeutics and vaccine formulations. The implementation of California COVIDNet allowed California to be a major global contributor of SARS-CoV-2 WGS data generating about 5% of all SARS-CoV-2 sequences in GISAID. Here, we describe the establishment of the IGED, an integrated database of SARS-CoV-2 sequences linked to individual patient epidemiologic data in a secured environment. To our knowledge, the IGED is a first of its kind database that enabled CDPH to develop genomic epidemiology tools (https://calcat.covid19.ca.gov/cacovidmodels/, https://pathogengenomics.ucsc.edu/tools) and monitor epidemiologic and evolutionary trends of recombinants (12) and persistent infections (13). For other public health organizations developing genomic surveillance systems like the IGED, we recommend the following key considerations: 1) establishing uniform sequencing and incident identifiers for matching between WGS and epidemiology data, 2) improving subspecies genomic classifications, 3) securing consistent funding, 4) creating a positive incentive for data contributors, and 5) developing interdisciplinary workforce communications.

Combining both ELR and non-ELR data was a provisional fix during an emergency response until all laboratories could be fully onboarded to ELR. We recommend onboarding laboratories to a consistent set of LOINC codes with strict ELR formatting for similar databases in the future. Nonetheless, the data integration methods developed for the IGED provide a framework for other public health organizations facing similar reporting challenges. To combat future emerging pathogens, it is paramount to establish uniform data requirements and formatting to ensure interoperability of the numerous data sources contributing to genomic and epidemiological surveillance.

We recommend that public health organizations report standardized subspecies classifications via ELR, which for SARS-CoV-2 are Pango lineages. Such classifications have been instrumental to various uses of the IGED including lineage proportion modeling (28, 29) and investigations into concerning infections (13). The number of pathogens for which there is a standardized subspecies classification nomenclature is limited, but recent efforts have expanded Pangolin (10) or Pangolin-like nomenclature to other viruses (33) (https://github.com/influenza-clade-nomenclature; https://github.com/mpxv-lineages/lineage-designation?tab=readme-ov-file). It is also critical that unique sequence identifiers such as sample IDs, GISAID accession numbers, or NCBI accession numbers be reported as ELRs via HL7 message to public health departments. These unique identifiers link directly to raw sequence data or consensus sequences to facilitate detection of mutations and potential outbreaks, even in the absence of standardized classification nomenclature.

Adequate and consistent funding, trained personnel, and the incentive to report are important factors to consider for the continuation of a successful genomic surveillance program for any pathogen. Incentives to report can include exclusive access to sequence databases, dashboards, computational resources, and tools like UCSC Big Tree Investigator. Facilities contributing to genomic surveillance efforts could be considered for special grants or provided access to trainings and workshops. They could also receive regular reports outlining the success of the sequencing efforts and analysis from the sequencing results.

Finally, public health organizations should establish partnerships with other government agencies and academic institutions as well as with genomics experts, laboratorians, data scientists, epidemiologists, and bioinformaticians with unique expertise needed to create similar databases to the IGED in the event of the next pandemic. Fostering these relationships can streamline the development of these databases and improve their data quality which will ultimately contribute to a more rapid, data-driven public health response.

## Acknowledgments

We gratefully acknowledge all data contributors, i.e., the authors from the originating laboratories and the submitting laboratories who generated and shared genetic sequence data and metadata via GISAID (the material on which this report is based). We thank the California Association of Public Health Laboratory Directors, public health laboratorians, and California COVIDNet laboratories partners for their contributions to California COVIDNet. We thank the CDPH CalREDIE and CDPH Data Teams for their stewardship of COVID data throughout the pandemic emergency. We are grateful to John Bell, and the CDPH COVID Clinical and CDPH Epidemiology Teams for their valuable feedback. Lastly, we thank the international team of volunteers that identify and name new SARS-CoV-2 Pango lineages.

## Data Availability

The WGS data presented in this study can be found in online repositories. The names of the repository/repositories and accession number(s) can be found at: https://www.ncbi.nlm.nih.gov/, Bioproject number PRJNA750736.

## Author Contributions

<u>Conceptualization:</u> MP, WW, ES, BB

<u>Supervision:</u> MP, WW, JM, ER, DAW

<u>Consulting:</u> TL, EB, JB, CM

<u>Writing:</u> JE, RR, KG, RK, EC, DAW

<u>Investigation:</u> JE, RR, DB, EL

<u>Data acquisition:</u> EC, THB, LN, BB, EAS, JE

<u>Database design:</u> JE, RR, MP, KG, RK, LT, RS, VS

<u>Database management:</u> JE, RR, DB, CK, LT, KG, RS, VS

<u>Database technology:</u> KG, RK, LT, CK, RS, VS, ER

<u>Project Management:</u> SG, PA, CM, MP, DAW

<u>Review & Editing:</u> JE, RR, KG, RK, ER, DAW

## Dictionary and Abbreviations

Abbreviations & Dictionary: Description
California COVIDNet: A multi-sector collaborative effort to achieve large-scale genomic surveillance of SARS-CoV-2 across California to monitor the spread of variants within the state, to detect new and emerging variants, and to characterize outbreaks in congregate, workplace, and other settings
California COVIDNet Terra database: Database from collaborators within California COVIDNet who use Terra.bio platform to store and analyze results.
CDPH: California Department of Public Health
CalCAT: California COVID Assessment Tool
CalREDIE: California Reportable Disease Information Exchange: a secure system that CDPH has implemented for electronic disease reporting and surveillance
CSV: Comma Separated Values
COVID-19 Case Registry: SARS-CoV-2 laboratory results that have been reported to CDPH electronically or manually by laboratories, healthcare providers, and local health departments
COVID-19 Hospitalization Registry: Dataset reported to CDPH following an All Facilities Letter (AFL) that required all hospitals in California to report specific patient-level information for each hospitalized patient who tested positive for COVID-19 on 27 July 2021 or later
ELR: Electronic Laboratory Reports
GISAID: Global Initiative on Sharing All Influenza Data: An international public repository for Influenza and SARS-CoV-2 sequencing data
HL7: Health Level Seven: International standards for the exchange of electronic health information between different healthcare systems
IGED: Integrated Genomic Epidemiology Database with a person-based deduplication scheme
IGED-SEQ: Integrated Genomic Epidemiology Database with a sequence-based deduplication scheme
LA: Los Angeles County
LIMS: Laboratory Information Management System
LOINC: Logical Observation Identifiers Names and Codes
Pango Lineage: The Pango lineage subspecies classification system is widely recognized as the standard for determining lineages of SARS-CoV-2
PAUI: Patient Anonymized Unique Identifier
PHI: Protected Health Information
PHL(s): Public Health Laboratory(ies)
PII: Personally Identifiable Information
RBAC: Role-Based Access Control
RPHO: Regional Public Health Officer
SD: San Diego County
SFTP: Secure File Transfer Protocol
SNOMED: Systematized Nomenclature of Medicine – Clinical Terminology
Snowflake: A cloud-based data platform that can store and analyze data
SQL: Structured Query Language
TAT: Turn-Around Time
Title 17: California Code of Regulations Title 17 refers specifically to SARS-CoV-2 sequence data required to be reported to CDPH per the updated Title 17, Section 2505, subsection (q) regulation
UCSC: University of California Santa Cruz
Vaccine Registry: Database that includes vaccine information reported to the California Immunization Registry.
VRDL: Viral and Rickettsial Disease Laboratory
WGS: Whole Genome Sequencing
WHO: World Health Organization

---

**S1 Table.** Two-tailed Spearman correlation test coefficients and p-values comparing the ranks of major lineage proportions between the IGED and GISAID sequences in California by quarter from Q1 2020 through Q1 2025. * **p** <0.05, ** **p** < 0.01, *** **p** < 0.01

